# The State-Level Association Between Safe Haven Laws and Rates of Infant Mortality in the United States

**DOI:** 10.1101/2022.11.16.22282430

**Authors:** Kathryn A. Thomas, Chloe J. Kaminsky

## Abstract

**Context:** Although the United States has safe haven laws, which allow mothers to safely relinquish their babies to designated safety points, rates of infant mortality remain significantly higher in the United States than other similarly developed countries. The current study is seeking to explore the state-level association between safe haven laws and infant mortality in the United States utilizing a legal epidemiological approach.

**Methods:** Several sources of publicly available data were combined to examine the state-level association between safe haven laws and rates of infant mortality. A backward stepwise regression was used to determine whether certain safe haven laws significantly predicted rates of infant mortality, while controlling for demographic variables.

**Findings:** After controlling for demographic variables including rates of health insurance and poverty, safe haven laws stating that only the mother can relinquish a child, laws that protect parents from criminal liability, and laws requiring the provider to provide legal information and referrals significantly predicted rates of infant mortality, with the total model accounting for 70.1% of variance in infant mortality.

**Conclusions:** Several elements of safe haven laws significantly predicted higher rates of infant mortality, including laws stating that only the mother may relinquish the child, laws that do not protect parents from criminal liability, and laws requiring a provider to provide legal information and referrals. These results have important implications for policymakers considering the reform of the safe haven laws. It is especially important to evaluate the effectiveness and origins of safe haven laws in the wake of the overturn of *Roe v. Wade*. Future studies should longitudinally examine how changes in safe haven laws lead to changes in rates of infant mortality.

**Policy Points:** ⍰ This is the first study to use a legal epidemiological approach to examine the state-level impact of specific elements of safe haven laws on rates of infant mortality.

⍰ Several elements of safe haven laws significantly predicted higher rates of infant mortality, including laws stating that only the mother may relinquish the child, laws that do not protect parents from criminal liability, and laws requiring a provider to provide legal information and referrals. These results have important implications for policymakers considering the reform of safe haven laws, which is especially important in the wake of the overturn of *Roe v. Wade*.

## Introduction

Rates of infant mortality remain significantly higher in the United States than in other similarly developed countries.^1^ In fact, infant mortality rates in the United States are three-to-four times higher than Hong Kong, Singapore, and Finland, and 31 to 51% higher than Canada and England.^2^ Although rates of infant mortality have decreased in recent decades, the United States’ decline in infant mortality has lagged behind other similarly developed countries, such that the United States ranked 12^th^ lowest in infant mortality worldwide in 1960 and 31^st^ in 2015.^2^ Research has shown that racial disparities in infant mortality in the United States have persisted for over a century, such that mortality rates for Black, Latinx, and Native American/Alaska Native infants remain significantly higher than rates for White and Asian infants.^2^ In fact, rates of infant mortality in the first 27 days following birth were 151% higher for Black infants than their White counterparts in 2017.^2^ Among the leading causes of infant mortality are congenital malformations, complications associated with low birthweight and short gestation periods, maternal complications, sudden infant death syndrome, and unintentional injuries.^3^

While some of these complications may be unavoidable, the high rate of infant mortality may be reflective of a larger policy problem regarding maternal and infant welfare in the United States. In reaction to the high rates of infant mortality and highly publicized cases of infanticide, all 50 states have passed various versions of safe haven laws, which provide a place for infants to be relinquished safely. In 1999, Texas passed the first safe haven law, with 15 states adopting safe haven laws in 2000 and all 50 states by 2008.^4^ Safe haven laws were originally enacted to address infant abandonment and provide mothers with a safe method of relinquishing their infants, with the ultimate objective of preserving infant health and preventing infanticide.^5^ Safe haven laws vary by state in terms of who may relinquish the infant, where the infant may be relinquished, and how old the infant is at the time of relinquishment.

Some state safe haven statutes protect certain parties, including medical providers and/or parents, from potential liability, while other states do not protect parties from potential criminal and civil liability, thus criminalizing those parties if relinquishment is not conducted according to the statutory requirements.^5^ This aspect of criminalization has important implications for the relationship between motherhood and the law. Some scholars argue that laws regarding child and maternal welfare may stigmatize mothers who deviate from the societal “motherhood” norm.^6^ Research has shown that laws that criminalize prenatal substance use tend to have negative effects on the mother’s health and decrease the likelihood of the mother seeking assistance.^7^ Further research is needed to understand the impact of safe haven policies that criminalize mothers on the health of mothers and their children.

For years, legal scholars have offered critiques of safe haven laws, arguing that safe haven laws were an emotional response to high-profile cases of infanticide and were not properly researched before being enacted, and, therefore, do not address the root problems of infant abandonment and infant mortality.^8^ Legal scholars have argued that safe haven laws may make citizens feel like crime and infant harm is being prevented, rather than offering concrete, evidence-based solutions.^9^ Moreover, safe haven laws have been criticized for ignoring racial, cultural, and socioeconomic issues that impact women’s reasons for relinquishment, as well as the complex needs of mothers during their pregnancy.^10^

Although a goal of safe haven laws is to ensure the safety of infants and reduce infant mortality, few studies have analyzed the relationship between safe haven laws and infant mortality. There are no national datasets aimed at understanding the use of safe havens in the United States, as most states do not mandate data collection on the number of infants surrendered to safe havens.^4^ Worldwide, there has been a call for more robust, long term data collection to understand the use of safe haven laws and the impact of safe haven laws on rates of infant mortality. ^4^ In one of the few studies examining the effectiveness of safe haven laws, Wilson and colleagues compared infant homicide rates in the United States from 1989-1998 and 2008-2017 to determine whether rates had changed after the enactment of safe haven laws across the United States; results revealed a 66.7% decrease in infant homicides from 1989-1998 to 2008-2017.^11^ The authors also examined whether the age limit of legal relinquishment predicted rates of infant homicide, and found that there was not a significant association between age limit of legal relinquishment and rates of infant homicide.^11^ The Centers for Disease Control and Prevention encouraged states to evaluate the effectiveness of safe haven laws in preventing infant homicides, and to consider other programs and policies that could reduce rates of infant homicides by providing economic support, affordable childcare, and skill-building for young parents.^11^

As these laws vary significantly by state, research is needed to understand what elements of safe haven laws, if any, predict infant mortality, including who may relinquish the infant, where the infant may be relinquished, and how old the infant is at the time of relinquishment. We utilize a legal epidemiological approach to examine the state-level association between specific elements of safe haven laws and rates of infant mortality in the United States, adding to the literature by including elements of safe haven laws beyond just those that establish age limits.

## Methods

### Procedure

Publicly available state-level data was combined from several data sources including: 1) Children Welfare Information Gateway’s List of State Statutes of Infant Safe Haven Laws as of 2021;^5^ 2) Center for Disease Control and Prevention state-level data on infant mortality rates per 100,000 live births in the United States in 2021;^12^ and 3) U.S. Census state-level estimates of race, poverty levels, percent of state that is rural, and health insurance rates from 2010 to 2019.^13^ Elements of safe haven laws included in the initial analyses were: 1) whether there is a right to reclaim the infant; 2) whether the provider must provide legal information and referrals; 3) the number of days after birth that an infant can be relinquished; 4) whether either parents may relinquish the infant; 5) whether only the mother may relinquish the infant; 6) whether an agent of the parent may relinquish the infant; 7) whether someone with legal custody can relinquish the infant; 8) whether the law specifies who can relinquish the infant; 9) whether the infant can be relinquished to a hospital; 10) whether the infant can be relinquished to a fire department; 11) whether the infant can be relinquished to law enforcement; 12) whether the infant can be relinquished to a church; 13) whether there is immunity from liability for providers; 14) whether parents can remain anonymous; 15) whether parents are protected from criminal liability 16) whether the provider must ask for medical information when an infant is relinquished. Data from all 50 states were included in the analyses. All variables were continuous, with the exception of the state-level coding of safe haven laws, which were coded 1 for the presence of the law and 0 for the lack of the law.

## Data Analysis

A hierarchical multiple regression was performed to examine the state-level association between safe haven laws and infant mortality. A backward stepwise approach was utilized to select the most salient correlate variables to control for in the first step of the regressions while reducing the likelihood of overfitting the model with variables not contributing to the variance in infant mortality. The backwards stepwise regression began with a full, saturated model that included all potential demographic variables informed by extant literature. Next, variables that contributed the least amount of variance were removed one-by-one to find a reduced model that best explained the data. The threshold for removing variables was *p* = .10.

## Results

The backwards stepwise approach revealed a final regression model which controlled for state-level percent of the population that falls below the poverty line, percent of the population with no health insurance, and percent of the population that has Medicaid insurance, which accounted for 53.2% of variance in infant mortality. See Table 1. In order to determine which legal variables to include in the second step of the model, another backwards stepwise approach was used, which revealed a final regression model that included presence of a law in which parents remain anonymous when relinquishing a child to a safe haven (*p* = .082), presence of a law in which only the mother may relinquish the child (*p* = .005), presence of a law in which either parent may relinquish the child (*p* = .094), presence of a law in which parents are protected from criminal liability (*p* = .018), presence of a law in which the provider must provide legal information and referrals (*p* = .024), and the number of days after birth that a child can be relinquished at a safe haven (*p* = .088). Thus, the second step of the model (including percent of the population that falls below the poverty line (*β* = 1.136), percent of the population with no health insurance (*β* = -.344), and percent of the population that has Medicaid insurance (*β* = -.767), presence of a law in which parents remain anonymous when relinquishing a child to a safe haven (*β* = .169), presence of a law in which only the mother may relinquish the child (*β* = .318), presence of a law in which either parent may relinquish the child (*β* = .182), presence of a law in which parents are protected from criminal liability (*β* = -.236), presence of a law in which the provider must provide legal information and referrals (*β* = .241), and the number of days after birth that a child can be relinquished (*β* = -.173)) predicted 70.1% of variance in infant mortality. See Table 2 for a list of states with each safe haven law that was significant in our analysis. The addition of the legal variables in the second step of the model led to a significant change in *R*^*2*^ (*F*(9,40) = 10.44, p < .001).

**Table 1.** Regression analyses examining the impact of safe haven laws on infant mortality.

| Predictors | *B* | *R*<sup>2</sup> |
| --- | --- | --- |
| *Step 1:* |  |  |
| Percent Below Poverty Line | 1.074*** | .532*** |
| Percent with No Health Insurance | -.305* |  |
| Percent with Medicaid | -.6.83*** |  |
| *Step 2:* |  |  |
| Percent Below Poverty Line | 1.136*** | .701*** |
| Percent with No Health Insurance | -.344* |  |
| Percent with Medicaid | -.767*** |  |
| Safe Haven Law: Number of Days after Birth that Child May Be Relinquished | -.173 |  |
| Safe Haven Law: Parents Remain Anonymous | .169 |  |
| Safe Haven Law: Parents are Protected from Criminal Liability | -.236* |  |
| Safe Haven Law: Only Mother May Relinquish | .218** |  |
| Safe Haven Law: Either Parent May Relinquish | .182 |  |
| Safe Haven Law – Provider Must Provide Legal Information and Referrals | .241* |  |

\*\*\*p < .001, \*\* p < .01, \* p < .05, N = 50.

**Table 2.** Safe Haven Laws by State.

| Type of Law | States with that Law |
| --- | --- |
| Law that Parents are Protected from Criminal Liability | AK, AZ, CA, CT, FL, GA, HI, ID, IL, IA, KS, KY, LA, MD, MA, MN, MO, MT, NE, NV, NM, NC, ND, OH, OK, PA, RI, SC, SD, TN, TX, VT, WA, WI |
| Law that Only Mother May Relinquish Child | GA, MD, MN, TN |
| Law that Provider Must Provide Legal Information and Referrals | CT, DC, DE, HI, IL, KY, LA, MI, MT, NM, ND, OH, OK, RI, SC, TN, WA, WI |

## Discussion

Rates of infant mortality are significantly higher in the United States than in other similarly developed countries. In fact, the United States ranks 26 out of 29 Organization of Economic Cooperation and Development (OECD) nations for rates of infant mortality.^1^ While safe haven laws were created to reduce cases of infanticide and other infant deaths,^4^ there has been no research that has examined the impact of the specific elements of safe haven laws (i.e., who may relinquish the infant, where the infant may be relinquished, and how old the infant is at the time of relinquishment) on rates of infant mortality to our knowledge. Thus, the goal of our study was to examine the state-level association between specific elements of safe haven laws and rates of infant mortality.

Results revealed that, when controlling for percent of the state’s population with no health insurance, percent of the population with Medicaid insurance, and percent of the population that falls below the poverty line, the presence of 3 elements of state safe haven laws were significant predictors of infant mortality, with the full model predicting 70.1% of the variance in infant mortality. Utilizing a backwards stepwise approach allowed for the inclusion of 16 elements of state-level safe haven laws in the analysis, and revealed that the elements of the safe haven law that most significantly predicted infant mortality were laws regarding who is permitted to relinquish a child, parental criminal liability, and whether the medical provider must provide legal information and legal referrals, rather than where the infant can be relinquished.

Specifically, results revealed that states with laws stating that only the mother may relinquish, that do not protect parents from criminal liability, and that require medical providers to provide legal information and legal referrals had higher rates of infant mortality.

Further research is needed to understand the mechanism by which these elements of safe haven laws are related to higher rates of infant mortality. One possible explanation is that laws that limit who may relinquish a child and laws that do not provide protection from criminal liability deter parents from relinquishing their infants. Future research should explore the mechanism by which these laws may be barriers to the use of safe havens. Specifically, no research has examined the impacts of criminalization on utilization of safe havens, although research has revealed the negative impacts of criminalizing prenatal substance use.^7,14^ It is possible that the criminalization of parents who use safe haven laws may deter mothers from safely relinquishing their infants, leading to increased rates of infant mortality, but further research is needed to explore the relationship.

Results of the current study also revealed that states with laws requiring that healthcare providers provide referrals and legal information to parent(s) utilizing safe havens had higher rates of infant mortality. One possible explanation of this relationship is the vast differences in the types of referrals and legal information required by each state statute, with some states offering substantial counseling and case management (WA Rev. Code § 13.34.360), and others only providing a pamphlet for the parent(s) (CT Gen. Stat. §§ 17a-58; 17a-59). Differences in the types of referrals and information offered may contribute to differing rates of mortality in the corresponding state. Future studies should explore the impact of specific types of referrals and information.

Not only do researchers need to explore these barriers in the law, but also policymakers. There is a need for more systematic data collection about the use of safe havens, and policymakers can advocate for laws requiring the collection of data concerning how often safe havens are used and by whom. The systematic collection of data would also allow researchers to examine potential disparities in the use of safe havens, as well as barriers that might hinder their use. Furthermore, systematic data collection could include data on the implementation of safe haven laws, which would allow for research to examine potential gaps in the law and current practices.

Legal scholars have identified the lack of education and resources to promote awareness of safe haven laws as a key issue in the effectiveness of safe haven laws in the United States.^4^ Mothers may be more likely to relinquish infants if they knew that these laws existed, but public education has been practically nonexistent and women are often unaware of the laws or how to use a safe haven.^15,16^ Research has revealed that mothers are most likely to seek information from health professionals and online health resources, and that 90% of expectant mothers preferred to consult more than one information resource before making a decision.^17^ Future studies can examine the role of education and promoting awareness of safe haven laws on the use of safe havens and the effectiveness of safe haven laws in reducing rates of infant mortality and infant homicide. It is especially important to consider who is utilizing safe haven laws in order to more effectively disseminate information about these laws. Additionally, other studies have suggested that mothers who illegally abandon or kill babies are on average 19 years old, single, no longer involved with the baby’s father, and concealed their pregnancy.^16^ Policies can then focus on disseminating information about safe havens to these populations who are most vulnerable to infant mortality.

Understanding the impact of state haven laws and their policy implications are especially important in recent months, as safe havens have been suggested, by Supreme Court Justice Amy Coney Barrett in the oral arguments in *Dobbs v. Jackson Women’s Health Organization*, as a way of alleviating the “burdens of parenting” and “obligations of motherhood” that will likely increase in wake of *Roe v. Wade* being overturned. It is evident that the connection between safe haven laws and abortion has been used in the conversation surrounding reproductive rights, making it even more important that safe haven laws are understood by researchers and policymakers alike. Safe haven laws were originally adopted with the purpose of reducing infant abandonment and infant homicide; they were not adopted to replace abortions or serve as an alternate option for women who cannot access abortion.^18^ In fact, women who are unable to get an abortion usually will not utilize safe havens or elect to place their infants up for adoption.^19^ While there is a need for future research that examines the effectiveness of safe haven laws, it is important to note that safe haven laws are only one piece of a larger puzzle that must be considered when considering women’s choices and reproductive justice.

While these results provide an important preliminary look at the association between state-level safe haven laws and rates of infant mortality, there are limitations to the study’s cross-sectional design. Future studies should utilize a longitudinal approach to determine how changes in safe haven laws lead to changes in infant mortality. Additionally, this study examines state-level trends rather than the individual impact of safe haven laws on parents’ decisions to utilize safe haven services. Future studies can examine individuals’ experiences utilizing safe havens and the barriers which may prevent their use on an individual level. The use of state-level data also limits the statistical power for analyses, and thus results may not capture the full impact of the safe haven laws on infant mortality, especially for laws that are enacted in few states.

## Data Availability

All data produced are available online.

